# Extended Validation of Transport Conditions of Thawed PF24 Units

**DOI:** 10.64898/2026.08.12.26360319

**Authors:** Dmitry Zlobin, Martin Jerez, Farrah Roberts, Jennifer Miller, Maria Proytcheva, Debra Smith, Yigit Baykara

**Affiliations:** Department of Pathology, University of Arizona College of Medicine, Tucson, Arizona, USA

## Abstract

**BACKGROUND:** The transport and storage conditions of thawed plasma are not strictly regulated by the FDA and applying red blood cell transport standard of 1–10°C to recently thawed plasma often results in high discard rates. This study evaluated the extended 120-hour (5-day) coagulation factor stability and sterility of thawed plasma frozen within 24 hours (PF24) following a 6-hour transport cooler simulation.

**STUDY DESIGN AND METHODS:** Fourteen PF24 units (8 group O, 6 group B) were thawed at 30–37°C and assigned as control (n=7, direct 1–6°C refrigeration) or experiment (n=7) units. Experiment units were held at room temperature for 30 minutes, stored in validated transport coolers for 6 hours, and then transferred to 1–6°C refrigeration. Measurements of temperature, prothrombin time (PT), Factor V (FV) activity, and Factor VIII (FVIII) activity were conducted at 0-, 6-, 24-, and 120-hour post-thaw. Sterility testing was performed at 0-hour and 120-hour using automated aerobic and anaerobic blood cultures.

**RESULTS:** No statistically significant differences were observed between control and experiment units at 120-hour for mean PT (15.09 vs. 15.16 seconds, *p* = .44), FV activity (81.14 vs. 74.57%, *p* = .23), or FVIII activity (61.86 vs. 53.00%, *p* = .22). Delta analysis (Δ_120h-0h_) confirmed equivalent factor decay rates between groups. All bacterial cultures showed no growth at 120-hour.

**CONCLUSION:** A 6-hour cooler time of thawed PF24 does not accelerate coagulation factor degradation or compromise sterility over an extended 5-day shelf life. These findings validate flexible inventory return policies, allowing blood banks to reduce product waste.

## 1 INTRODUCTION

Preserving the quality of blood products during storage and transport is critical to ensure the safety and efficacy of transfusion. Thawed fresh frozen plasma (FFP) and plasma frozen within 24 hours (PF24) are highly sensitive to temperature changes after thawing, as the functional activity of critical, heat-labile coagulation factors such as Factor V (FV) and Factor VIII (FVIII) and factors with a short half-life such as Factor VII gradually degrades over time.^1–3^ The transport and storage conditions of thawed plasma products are not specifically regulated by the U.S. Food and Drug Administration; AABB Standard 5.1.8A specifies a transport temperature range of 1–10°C for refrigerated blood products. Although this standard was developed primarily for red blood cells, many institutions apply it to thawed plasma products by convention in the absence of a plasma-specific transport guideline.^4,5^ The 1–10°C temperature range is often difficult to maintain during real-world clinical practices such as an immediate increase in temperature due to thawing procedure itself or routine transportation, leading to high rates of product discard.^6^

We previously demonstrated that short-term (up to 6 hours) temporary storage of thawed FFP within validated transport coolers does not statistically compromise FV activity, FVIII activity, or prothrombin time (PT).^1^ While those findings supported institutional policy revisions allowing the safe return of cooler-stored thawed FFP back to inventory, that initial evaluation was restricted to a brief 6-hour post-thaw window and utilized exclusively group B plasma units. Importantly, the long-term impacts of delayed cooling on extended 5-day storage coagulation parameters remain unaddressed. Furthermore, potential concerns regarding delayed bacterial growth following an initial ambient temperature exposure were not experimentally monitored.

To address these limitations, this expanded study evaluates the extended coagulation factor stability and sterility of thawed PF24 units subjected to simulated cooler transport over a 120-hour (5-day) post-thaw period. By incorporating both group O and group B plasma units, alongside rigorous microbiological surveillance, we seek to comprehensively validate extended preservation, thereby facilitating improvements in plasma product inventory management and reducing unnecessary product waste.

## 2 METHODS

### 2.1 Unit Preparation and Thawing

A total of 14 PF24 units—comprising 8 group O units and 6 group B units—were utilized. The units were randomly allocated as control units (n=7; 4 group O, 3 group B) and experiment units (n=7; 4 group O, 3 group B). Prior to the initiation of the experiment, all units were maintained under standard temperature conditions in a Helmer i.Series® Upright Plasma Freezer (Helmer Scientific, Noblesville, IN) at –30°C (referred to hereafter as freezer). All units were thawed utilizing the QuickThaw Plasma Thawing System (Helmer Scientific, Noblesville, IN) at 30–37°C according to established institutional protocol.

### 2.2 Simulated Transport and Storage Conditions

Immediately following the thawing process, all 7 control units were placed directly into a Helmer i.Series® Blood Bank Refrigerator (Helmer Scientific, Noblesville, IN) set to ideal storage temperatures of 1–6°C (referred to hereafter as refrigerator). To simulate routine clinical handling and transport delays, the 7 experiment units were first held on an open laboratory bench at ambient room temperature for exactly 30 minutes.

Following this ambient room exposure, the units were distributed into two validated transport coolers. The coolers are Igloo Playmate MaxCold Cooler Series (Igloo, Toronto, Canada) validated to maintain a temperature of 1–10°C for up to 6 hours, containing one refrigerated and two frozen cooling bricks (Coleman Brite Ice™, The Coleman Company, Wichita, KS). One of the coolers housed 1 group O unit and 2 group B units, and the other cooler housed 3 group O units and 1 group B unit. The units remained inside the insulated coolers for a continuous duration of 6 hours. Immediately upon reaching the 6-hour time point, all experiment units were transferred into a refrigerator for the remainder of the 120-hour study period.

### 2.3 Sampling Protocol and Thermal Surveillance

To evaluate potential bacterial contamination risks secondary to the initial room temperature transport simulation, dedicated sterility testing was performed on all units at 0-hour (baseline) and 120-hour (5-day) sampling intervals. Surface temperature measurement and coagulation sampling of all units were conducted at designated post-thaw intervals: 0-hour (baseline), 6-hour, 24-hour, and 5-day (120-hour). At each time point, individual units were removed from their respective storage environments one at a time for processing. To prevent environmental thermal contamination, total cumulative exposure outside of the refrigerator or cooler was strictly limited to a maximum of 15 minutes at each interval. The units were spiked using a sampling site coupler (Terumo, Tokyo, Japan) and a 21 Gauge BD Vacutainer® Eclipse™ Blood Collection Needle (Becton Dickinson, Franklin Lakes, NJ) was used for each distinct sampling event to maintain closed-system integrity. Surface unit temperatures were sequentially recorded using a calibrated Traceable® Infrared Thermometer Gun (Thomas Scientific, Swedesboro, NJ) at every interval. Approximately 10 mL aliquots were extracted from each unit for every time point and split evenly into three 6 mL BD Vacutainer® Serum Tubes (Becton Dickinson, Franklin Lakes, NJ) for PT, FV activity and FVIII activity testing. All harvested coagulation vacutainers were transferred immediately to a freezer to preserve coagulation factor activity. Alongside the coagulation samples, an additional 20 mL aliquot was extracted from each experiment and control unit. This volume was inoculated evenly into automated blood culture bottles: 10 mL into BD BACTEC™ Plus Aerobic/F Culture Vials and 10 mL into BD BACTEC™ Lytic/10 Anaerobic/F Culture Vials (Becton Dickinson, Franklin Lakes, NJ). The study design and experimental procedures are demonstrated in Figure 1.

### 2.4 Coagulation Analysis

Frozen samples were batched and transported to the special coagulation laboratory inside Styrofoam containers packed with dry ice. Quantitative analyses for FV functional activity (%), FVIII functional activity (%), and PT (seconds) were performed on ACL TOP 550 CTS Coagulation Analyzer (Instrumentation Laboratory Werfen, Barcelona, Spain).

### 2.5 Microbiological Surveillance

Inoculated culture pairs were transferred immediately to the clinical microbiology laboratory for continuous automated incubation and growth monitoring in BD BACTEC™ FX blood culture system (Becton Dickinson, Franklin Lakes, NJ).

### 2.6 Data analysis

All data were collected and analyzed using Microsoft®Excel® for Microsoft 365 MSO. We used one-tailed t-test to compare the means of two groups on single independent variables such as temperature, PT, FV activity, and FVIII activity at time points 0-hour, 6-hour, 24-hour, and 120-hour. We also used one-tailed t-test for comparison of change (delta analysis) in the means of the same variables between time points 120-hour and 0-hour. A p-value of <.05 was considered statistically significant.

## 3 RESULTS

The mean temperature of PF24 units measured immediately after thawing was not different between the control and experiment units (30.26 vs. 29.36°C, *p* = .23). At 6-hour, the control units kept in the refrigerator were significantly colder than the experiment units which were held for 30 min on the bench and then placed in a standard transport cooler (4.26 vs. 10.34°C, *p* < .001). This significant difference in temperature disappeared at 24-hour storage (3.66 vs. 3.86°C, *p* = .17). There was a statistical difference in the mean temperatures between the control and experiment units at 120-hour (4.46 vs. 3.80°C, *p* < .001). Moreover, the mean volume of the control units was slightly less than that of the experiment units (231.86 vs. 276.14 mL, *p* = .047).

The mean PT of the samples collected immediately after thawing was not different between the control and experiment units (13.11 vs. 13.20 seconds, *p* = .41). The differences between the mean PT of the control and experiment units were not significant at 6-hour (13.26 vs. 13.06 seconds, *p* = .28), 24-hour (13.76 vs. 13.71 seconds, *p* = .46), and 120-hour (15.09 vs. 15.16 seconds, *p* = .44).

The mean FV activity of the samples collected immediately after thawing was not different between the control and experiment units (93.57 vs. 90.43%, *p* = .37). The lack of statistical difference persisted at 6-hour (93.71 vs. 89.43%, *p* = .32), 24-hour (91.00 vs. 83.86%, *p* = .22), and 120-hour (81.14 vs. 74.57%, *p* = .23).

The mean FVIII activity of the samples collected immediately after thawing was also not different between the control and experiment units (97.71 vs. 86.86%, *p* = .28). There was no statistical difference also at 6-hour (86.14 vs. 79.86%, *p* = .35), 24-hour (73.86 vs. 67.71%, *p* = .34), and 120-hour (61.86 vs. 53.00%, *p* = .22). These results are summarized in Table 1.

**Table 1.** Mean temperature, prothrombin time, Factor V and Factor VIII activities of samples taken at different time points from the units. Temp, Temperature; PT, Prothrombin Time; SD, standard deviation.

| Units | Time (h) | Temp (C°) (mean ± SD) | PT (s) (mean ± SD) | FV% (mean ± SD) | FVIII% (mean ± SD) |
| --- | --- | --- | --- | --- | --- |
| Experiment | 0 | 29.36 ± 1.79 | 13.20 ± 0.58 | 90.43 ± 16.01 | 86.86 ± 32.53 |
| Control | 0 | 30.26 ± 2.52 | 13.11 ± 0.78 | 93.57 ± 18.57 | 97.71 ± 36.71 |
| Experiment | 6 | 10.34 ± 0.96 | 13.06 ± 0.47 | 89.43 ± 13.73 | 79.86 ± 29.03 |
| Control | 6 | 4.26 ± 0.52 | 13.26 ± 0.76 | 93.71 ± 18.44 | 86.14 ± 31.87 |
| Experiment | 24 | 3.86 ± 0.45 | 13.71 ± 0.61 | 83.86 ± 15.07 | 67.71 ± 26.99 |
| Control | 24 | 3.66 ± 0.26 | 13.76 ± 0.86 | 91.00 ± 18.75 | 73.86 ± 28.38 |
| Experiment | 120 | 3.80 ± 0.24 | 15.16 ± 0.80 | 74.57 ± 13.89 | 53.00 ± 19.37 |
| Control | 120 | 4.46 ± 0.18 | 15.09 ± 0.93 | 81.14 ± 18.15 | 61.86 ± 21.64 |

We also performed delta analysis to ensure that the lack of statistical difference between the means of coagulation parameters of control and experiment units were not due to lower baseline coagulation factor levels in the control units. Comparison of the mean changes in PT, FV activity, and FVIII activity between the time points 120-hour and 0-hour showed no statistical difference between the experiment and control units (Δ1.96 vs. Δ1.97, *p* = .48, Δ-15.86 vs. Δ-12.43, *p* = .22, and Δ-33.86 vs. Δ-35.86, *p* = .41, respectively).

Microbiological surveillance confirmed the maintenance of product sterility throughout the extended storage period. Automated baseline (0-hour) aerobic and anaerobic blood cultures from the experiment and control units demonstrated no bacterial growth. Following the 5-day storage period, repeat automated cultures at 120-hour similarly demonstrated a complete absence of microbial growth in both aerobic and anaerobic vials for all evaluated experiment and control units. No delayed bacterial proliferation or contamination was observed in any culture bottle.

## 4 DISCUSSION

Building upon the previous validation of short-term thawed FFP transport, this expanded study demonstrates that temporary storage in transport coolers up to 6 hours does not compromise the extended 5-day stability or sterility of thawed PF24 units. By tracking coagulation parameters and monitoring microbiological safety over a 120-hour storage window, these findings provide robust evidence to support flexible blood bank return policies, thereby minimizing unnecessary product discard without introducing safety risks.

The initial temperature measurements immediately post-thaw reflect the clinical reality of blood bank operations. Thawed FFP and PF24 units inherently retain substantial heat from the water bath and bringing units down to the ideal 1–6°C requires prolonged cooling. While the refrigerator-stored control units naturally cooled faster than the cooler-enclosed experiment units during the first 6 hours (*p* < .001), this initial lag in cooling did not translate into an accelerated decay of coagulation factors. By the 24-hour time point, the experiment units had successfully equilibrated to standard refrigeration temperatures, matching the control units for the remainder of the 5-day storage period. Although a statistically significant temperature difference re-emerged at 120-hour (3.80 vs. 4.46°C, *p* < .001), both values remained well within the 1–6°C storage range, and the direction of the difference—experiment units being slightly colder—argues against any clinically meaningful thermal compromise. This minor discrepancy is most likely attributable to brief refrigerator door openings and the slightly smaller mean volume of the control units (231.86 mL vs. 276.14 mL, *p* = .047), which makes them more susceptible to immediate ambient temperature changes due to a larger surface-area-to-volume ratio and a smaller thermal mass.

Our data revealed that the temporary thermal differences observed at 6-hour did not significantly accelerate the degradation of heat-labile coagulation factors. Factor V and Factor VIII activities, and overall PT values, which are a surrogate for Factor VII activities, were not statistically different between the control and experiment units at serial measurements up to 120 hours. While a gradual, expected decline in Factor VIII functional activity was noted in both cohorts over the 5-day storage period (∼97% to ∼61% in control units and ∼86% to ∼53% in experiment units), this mirrors well-documented baseline decay curves for thawed plasma and was not exacerbated by the temporary cooler storage.^2,7^ The preservation of factor levels above the critical hemostatic threshold of 30%^8^ over 5 days confirms that the initial delay in cooling to 1-6°C does not jeopardize the therapeutic efficacy of the product. Our findings also align with those of Lamboo et al., who observed no significant loss of FV activity or prolongation of PT in thawed FFP stored at room temperature for up to 6 hours, with only a modest 16% decline in FVIII activity over that period.^6^ Even this slight FVIII reduction is comparable to what we observed in the early post-thaw hours and was not exacerbated by cooler storage. Ramirez-Arcos et al. demonstrated that multiple short-term exposures of thawed plasma to room temperature did not significantly affect the stability of coagulation factor levels compared with continuously refrigerated controls over a 5-day storage period.^9^ Furthermore, our inclusion of both group O and group B plasma units adds an important layer of generalizability that the initial study lacked.^1^ Because baseline Factor VIII levels naturally vary by ABO blood group (with group O individuals typically exhibiting lower baseline levels), testing a mixed-group cohort ensures that our findings hold true across different intrinsic factor concentrations.

Beyond functional efficacy, a principal concern regarding temporary room-temperature and cooler exposure is the theoretical risk of promoting delayed bacterial growth. Our study directly addresses this biosafety concern through microbiological surveillance using automated aerobic and anaerobic blood cultures. The absence of microbial growth at both baseline and day 5 across all tested experiment units strongly indicates that the 30-minute ambient bench exposure followed by closed, insulated cooler storage maintains strict product sterility. Our results are consistent with those of Ramirez-Arcos et al., who performed deliberate inoculation studies in thawed plasma with four clinically relevant organisms (*Serratia liquefaciens, Pseudomonas putida, Staphylococcus epidermidis, and Pseudomonas aeruginosa*) and found that the duration of room-temperature exposure (30 vs. 60 minutes) did not result in statistically significant differences in bacterial concentrations; notably, *S. liquefaciens* proliferated during cold storage regardless of room-temperature exposure duration, while *S. epidermidis and P. aeruginosa* survived but did not grow, indicating that storage temperature rather than the delayed cooling is the dominant factor governing bacterial behavior in thawed plasma.^9^ Sheffield et al. similarly confirmed unit sterility through 120 hours of refrigerated post-thaw storage using an automated microbial detection system.^2^ The inherently low bacterial contamination rate of plasma products is further supported by the acellular nature of plasma, which lacks the platelet-derived metabolic substrates (such as lactic acid) that have been shown to enhance bacterial replication in platelet concentrates stored at room temperature.^10^

These findings have direct operational and regulatory implications for transfusion services. Because transport and return parameters for thawed FFP or PF24 are not strictly mandated by the U.S. Food and Drug Administration, institutions have the flexibility to establish internal, validated criteria for inventory management. Historically, applying the rigid 1–10°C RBC transport threshold to recently thawed FFP or PF24 has forced blood banks to discard perfectly viable units simply because they have not had sufficient time to cool down upon return. By demonstrating extended 5-day coagulation factor stability and uncompromised sterility following a 6-hour time in standard cooler, this study provides the formal validation necessary to enact robust inventory return policies. Implementing a standardized return protocol for freshly thawed FFP or PF24 can substantially reduce product waste and lower operational costs.

While these results strengthen the case for policy optimization, certain limitations must be acknowledged. First, although the sample size (n=14) was sufficient to demonstrate statistical equivalence across primary endpoints, it may possess limited statistical power to detect minute, subtle variations in specific coagulation sub-components. Second, this study was conducted within a single-center environment utilizing apheresis-derived PF24 units and specific validation cooler setups. Consequently, the absolute generalizability of these exact temperature curves to whole-blood-derived plasma or highly disparate transport configurations may vary. Finally, our functional assessment was restricted to PT, Factor V, and Factor VIII. While these parameters serve as excellent surrogates for overall plasma quality and labile factor preservation, they do not capture the entire spectrum of the coagulation cascade or in vivo clinical transfusion outcomes. Future multi-center studies utilizing broader coagulation panels may offer additional refinement. These limitations suggest that institutions with substantially different operational workflows should consider internal validation before adopting similar policies.

## 5 CONCLUSION

In conclusion, short-term exposure of thawed PF24 units to simulated transport conditions for up to 6 hours does not accelerate coagulation factor degradation or compromise sterility over an extended 5-day shelf life. These data provide comprehensive validation for blood bank services to confidently accept cooler-transported thawed FFP or PF24 units back into active inventory, achieving significant reductions in resource waste without compromising patient safety or clinical efficacy.

## Data Availability

All data produced in the present work are contained in the manuscript

## BULLETED STATEMENT

1. What’s already known about this topic? There is a previous study done and published in by the author Yigit Baykara regarding validation of transport conditions of thawed plasma units which showed that short-term storage of thawed fresh frozen plasma in validated transport coolers does not compromise coagulation factor activity or PT at least for up to 6 hours.
2. What does this study add? This study is an extended version of the previous study that addresses the limitations of the previous study and evaluates the plasma stability up to 5 days post-thaw (time period after it is returned back to the inventory and re-labeled as “thawed plasma”) after temporary storage in transport coolers in terms of factor activity, prothrombin time and sterility.

## AUTHOR CONTRIBUTIONS

Yigit Baykara contributed to all aspects of this research, including conceptualization, data collection, data analysis, conducting the literature review, and manuscript preparation. Dmitry Zlobin contributed to data collection and manuscript preparation. Martin Jerez, Farrah Roberts, Jennifer Miller contributed to conceptualization and data collection. Debra Smith and Maria Proytcheva contributed to conceptualization and manuscript preparation.

## ACKNOWLEDGMENTS

We thank Cassandra Simmons, Joanne Cavin, Lucy Sanford, Paul Brunstein, and Alastair Dunnett at Banner University Medical Center Tucson for their dedicated efforts and assistance with logistical planning and laboratory testing in this study.

## FUNDING INFORMATION

This study was funded by Banner University Medical Group Tucson, Department Incentive Funds.

## CONFLICT OF INTEREST STATEMENT

The authors have disclosed no conflicts of interest.

## PATIENT CONSENT STATEMENT

Patient consent is not applicable as no individual patient data is included in this manuscript.

## ETHICAL STATEMENT

This article does not contain any studies with human participants or animals performed by any of the authors. Therefore, ethical approval and informed consent were not required

## DATA AVAILABILITY STATEMENT

The data that support the findings of this study are available from the corresponding author upon reasonable request.

---

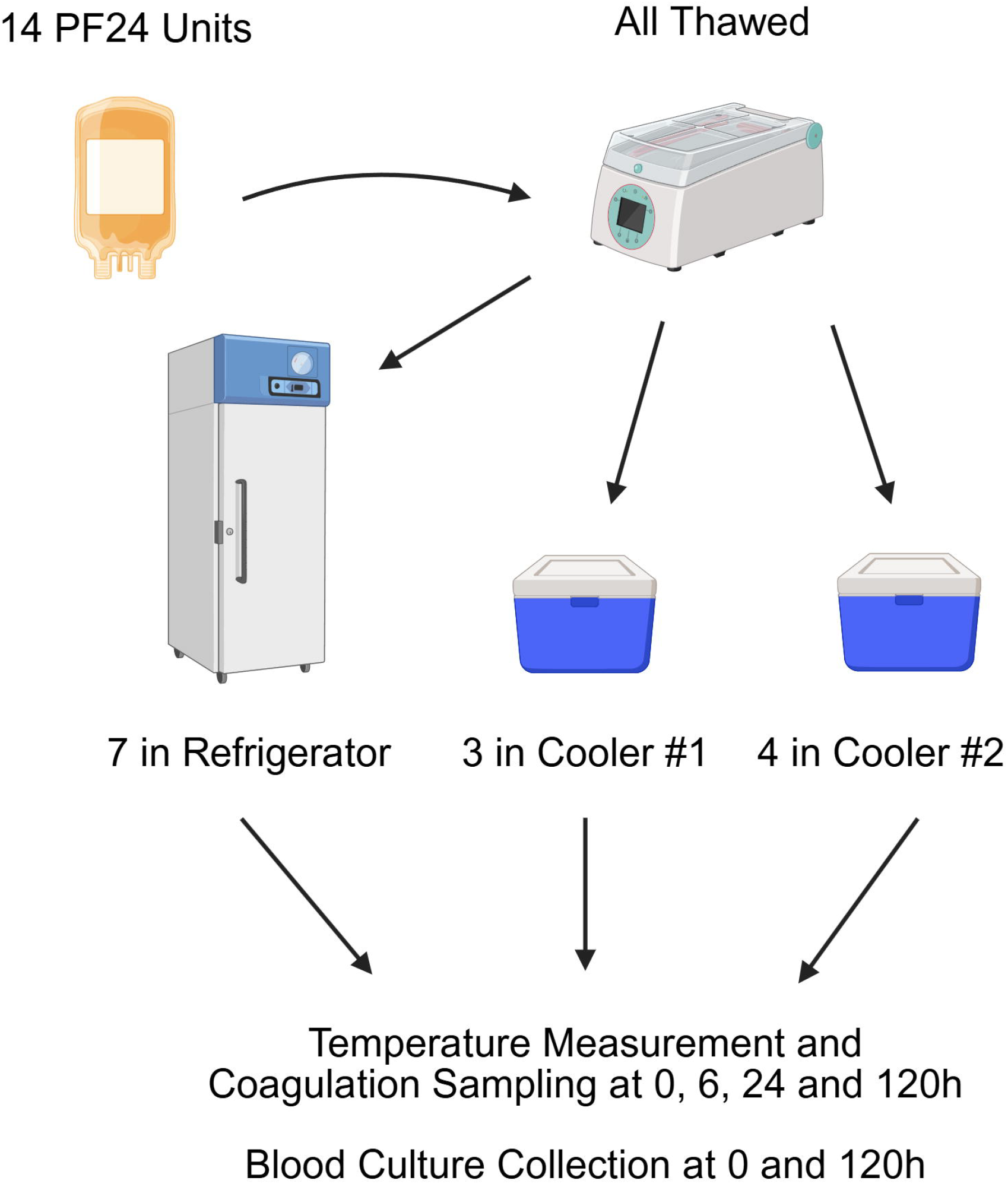

## Notes

### Competing Interest Statement

The authors have declared no competing interest.

### Author Declarations

Institutional Review Board of the University of Arizona waived ethical approval for this work.

